# Order-of-mutation effects on cancer progression: models for myeloproliferative neoplasm

**DOI:** 10.1101/2023.08.16.23294177

**Authors:** Yue Wang, Blerta Shtylla, Tom Chou

## Abstract

In some patients with myeloproliferative neoplasms (MPN), two genetic mutations are often found, JAK2 V617F and one in the TET2 gene. Whether or not one mutation is present will influence how the other subsequent mutation affects the regulation of gene expression. When both mutations are present, the order of their occurrence has been shown to influence disease progression and prognosis. We propose a nonlinear ordinary differential equation (ODE), Moran process, and Markov chain models to explain the non-additive and non-commutative mutation effects on recent clinical observations of gene expression patterns, proportions of cells with different mutations, and ages at diagnosis of MPN. These observations consistently shape our modeling framework. Our key proposal is that bistability in gene expression provides a natural explanation for many observed order-of-mutation effects. We also propose potential experimental measurements that can be used to confirm or refute predictions of our models.

## 1 Introduction

Different genetic mutations are found in patients with myeloproliferative neoplasm (MPN), a cancer of the bone marrow. These mutations are known to have different effects on cell behavior [30, 15, 27, 36]. In this paper, we focus on two common mutations in MPN, JAK2 V617F (henceforth abbreviated as JAK2) and TET2.

JAK2, the Janus kinase 2, mediates cytokine signaling to control blood cell proliferation, while the TET2 protein catalyzes oxidation of 5-methylcytosine, thereby epigenetically influencing expression of other genes. It has been shown *in vitro*, that JAK2 and TET2 mutations each confer a competitive growth advantage in some myeloid cells [4, 10]. Once the JAK2 or TET2 mutation appears in certain myeloid cells, such cells will have higher proliferation rates than myeloid cells without such mutations. This growth advantage is “modest” and might take years to manifest itself as an increased proportion of cells carrying these mutations in the total cell population. Thus, it is common to find cells in a patient with different numbers of mutation types. Moreover, in certain patients with *both* JAK2 and TET2 mutations, it is possible to infer which mutation appears first.

Ortmann et al. [40] reported that different mutational patterns (including the order of mutations) in hematopoietic cells and progenitor cells are related to differences in gene expression patterns, clonal evolution, and even macroscopic properties. Specifically, a mutation can differentially regulate gene expression by different amounts depending on whether or not another type of mutation preceded it. Therefore, the change in gene expression level when one mutation appears cannot simply be added. We call such phenomena “**non-additivity**”. Additionally, patients in which the JAK2 mutation appears before the TET2 mutation, have different gene expression levels, percentage of cells with only one mutation, and age at diagnosis than patients in which the TET2 mutation appears before the JAK2 mutation. This observation implies that the order of the first appearance of these two mutations matters. We describe such phenomena as “**non-commutative**”.

In Section 2, we summarize the clinical observations reported by Ortmann et al. [40]. In Section 3, we summarize previous models for such clinical observations [40, 49, 46, 24, 3, 12, 51, 50, 33, 52] and compare these models with our new models. We then build nonlinear ordinary differential equation (ODE) models to explain the observations regarding gene expression and list experimental evidence that supports our models in Section 4. In Section 5, we present a generalized Moran process model and three different mechanisms to explain the observations regarding clonal evolution and ages at diagnosis. We conclude with some discussion in Section 6. A more detailed review of previous models is given in Appendix A, while an alternative, but related Markov chain model to explain non-commutative effects of mutations on gene expression is presented in Appendix B.

## 2 Clinical observations on the effects mutation order

For patients exhibiting cells with *both* JAK2 and TET2 mutations, one might ask: Which mutation occurred first in the patient? If we find cells with only JAK2 mutations, cells with both JAK2 and TET2 mutations, but no cells with only the TET2 mutation, then the JAK2 mutation must have appeared in the patient before the TET2 mutation. Such patients are classified as JAK2-first. Patients in which we find doubly mutated cells and TET2-only cells but not JAK2-only cells are classified as TET2-first. If a patient carries JAK2-only cells, TET2-only cells, and JAK2-TET2 cells, then both JAK2 and TET2 mutations occurred independently in wild-type cells and more information, such as other associated mutations or tagging that resolves subpopulations, is needed to infer their temporal order of appearance. Such patients were not considered by Ortmann et al. [40]. For more complex samples that contain cells with multiple types of mutations, one can use different algorithms to determine the probabilities of different orders of mutations from sequencing data [14, 42, 44, 25, 17]. However, patients with ambiguous cell populations (JAK2-only cells, TET2-only cells, and JAK2-TET2 cells) were not considered by Ortmann et al. [40].

Besides inferring the order of mutations, Ortmann et al. [40] also measured bulk gene expression levels from MPN-patient-derived populations of cells containing different sets of mutations. Their observations are summarized in Table 1 in which *x*^∗^ denotes the steady state expression level of gene X in a cell and the subscripts define mutation status of the cell.

1. Some genes are up-regulated (or down-regulated) by a JAK2 mutation only if the TET2 mutation is not present. If the TET2 mutation is also present, the expression of these genes is not affected. Thus 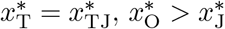 or 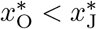.
2. Other genes are up-regulated (or down-regulated) by JAK2 mutations only if TET2 mutations are also present; but they are not affected if the TET2 mutation is *not* present. For these cases, 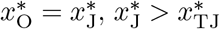 or 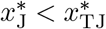.
3. Ten genes (AURKB, FHOD1, HTRA2, IDH2, MCM2, MCM4, MCM5, TK1, UQCRC1, WDR34) are up-regulated in cells with JAK2 mutations if TET2 mutations are not present, but they are down-regulated by JAK2 mutations if TET2 mutations *are* present. This scenario corresponds to 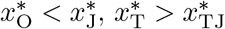.
4. Different orders of appearances of JAK2 and TET2 mutations seem to have different effects on other genes so that 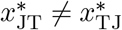. These conclusions are inferred from other indirect evidence (*e*.*g*., JAK2-first cells are more sensitive to ruxolitinib than TET2-first cells [40]). Observations **(1-3)** can be regarded as **non-additivity**, since the effect of JAK2 mutation differs with or without TET2 mutation. In other words, 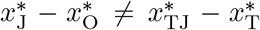. Observation **(4)** represents **non-commutativity** since exchanging the order of acquiring different mutations can lead to different expression levels or cell states [29]. Mathematically, 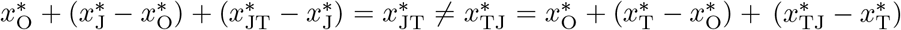. In fact, if the gene expression levels are additive with respect to multiple mutations, namely 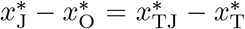 and 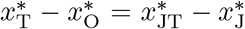, then it is also commutative: 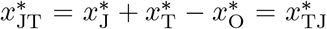. Therefore, non-commutativity is a special case of non-additivity. At the cell or tissue level, Ortmann et al. [40] also report two observations specifically related to non-commutativity:
5. In TET2-first patients, the percentage of cells with just one mutation (TET2) is significantly higher than the percentage of JAK2-only cells in JAK2-first patients.
6. At diagnosis, JAK2-first patients are significantly younger than TET2-first patients.

**Table 1:** Definition of stationary gene expression levels *x*^∗^ for cells with different mutation patterns.

|  | w/o JAK2 mutation | with JAK2 mutation |
| --- | --- | --- |
| w/o TET2 mutation | $x_O^*$ | $x_J^*$ |
| with TET2 mutation | $x_T^*$ | $x_{JT}^*$ (JAK2-first)<br>$x_{TJ}^*$ (TET2-first) |

Ortmann et al. [40] also report other observations such as differences in MPN classification and risk of thrombosis between JAK2-first and TET2-first patients. These are covered by observations **(1–6)**, particularly **(4)**, and we do not explicitly discuss them here.

## 3 Comparison between previous models and our models

There have been models put forth that explain observations **(1-6)**. We briefly summarize previous models in this section. See Appendix A for a detailed review of previous models. These past models and our set of models are compared in Table 2. Our models provide better coverage of the observed phenomena and can be concatenated for a more complete picture of MPN progression. We now provide an overview of our more complete analysis, filling in some mechanistic explanations of observations **(1-6)**.

**Table 2:** A summary of studies in mutational order and how they address observations given in [40]. Previous studies (top) and corresponding explanations are compared with the understanding afforded by our proposed mechanisms and models (bottom).

| **Previous Studies** | Key assumptions | Quantitative? | Observation(s) explained |
| --- | --- | --- | --- |
| Kent and Green [24] | JAK2 and TET2 mutations compete for the same regulation region | ✗ | (4) |
| Kent and Green [24] | JAK2 and TET2 have different effects on microenvironments | ✗ | (4) |
| Roquet et al. [46] | Mutations work like recombinases | ✗ | (4) |
| Clarke et al. [12]<br>Talarmain et al. [51, 50]<br>Mazaya et al. [33] | Gene expression satisfies a generalized boolean network model with multistability | ✓ | (4) |
| Ortmann et al. [40]<br>Ascolani and Liò [3]<br>Clarke et al. [12] | JAK2 and TET2 mutations confer different advantages to cell proliferation | ✓ | (5) |
| Teimouri and Kolomeisky [52] | JAK2 and TET2 mutations bring different advantages to cell proliferation | ✓ | (6) |
| **Current Analysis** | Key assumption | Quantitative? | Observation(s) explained |
| ODE model | Gene expression satisfies a nonlinear ODE with bistability | ✓ | (1, 2, 3, 4) |
| Markov chain model | Gene expression satisfies a Markov chain with bistability | ✓ | (4) |
| Moran process,<br>Mechanism (A) | JAK2 and TET2 mutations bring different advantages to cell proliferation | ✓ | (5, 6) |
| Moran process,<br>Mechanism (B) | Mutation rates for JAK2 and TET2 are different | ✓ | (5, 6) |
| Moran process,<br>Mechanism (C) | JAK2 mutation can induce TET2 mutation | ✓ | (5, 6) |

Observations **(1)** and **(2)**, the up-regulation of certain genes depending on the presence and absence of certain mutations, form a common “logic gate” in which expression levels can be changed if and only if both conditions are met. We first construct a simple nonlinear ODE model to explain observations **(1, 2)** by defining a threshold that is passed if and only if both conditions are satisfied. This model will serve as a building block for an explanation of observation **(3)**, for which no model has thusfar been proposed. We also explain **(3)** using a nonlinear ODE model and find two candidates for a hidden factor in this model. Some regulatory terms in our model have been verified experimentally, and we propose experiments to examine other regulatory relationships.

Kent and Green [24] explain the non-commutative order-of-mutation effects (observation **(4)**) by invoking hypothetical mechanisms that lack experimental evidence, while Roquet et al. [46] simply proposed a mathematical space in which operators are not commutative and not really connected to genetic mutations. Clarke et al. [12], Talarmain et al. [51, 50], and Mazaya et al. [33] all use (generalized) boolean networks to explain observation **(4)**. The state space is discrete, and the deterministic dynamics with many parameters are chosen artificially with little justification. At the single-cell level, gene expression levels are discrete and stochastic, but at the bulk level it is approximately deterministic and continuous.

Therefore, we propose a nonlinear ODE model (deterministic, continuous-state) and a Markov chain model (stochastic, discrete-state) to describe observation **(4)**.

Observation **(5)** can be simply explained by assuming that different mutations give rise to different proliferation advantages, as qualitatively described in Ortmann et al. [40]. Ascolani and Liò [3] model driver and passenger mutations using a similar assumption, but not for MPN.

The only possible explanation put forth for younger JAK2-first patients at diagnosis (observation **(6)**) was provided by Teimouri and Kolomeisky [52] who also assumed that the different mutations carry different proliferation advantages. In their model, the second mutation can appear if and only if the first mutation rapidly expands in the cell population. Because they assume that reaching the final state that all cells have both mutations is conditioned on no extinction once a mutation appears, they underestimate the predicted time to reach the final state.

To study observations **(5, 6)**, we consider a generalized Moran process, which is a more realistic model for describing the population dynamics of hematopoietic stem and progenitor cells. We find that three different parameter limits, describing three distinct biological mechanisms (including that proposed by Teimouri and Kolomeisky) can reproduce observations **(5, 6)** separately.

## 4 Models for non-additivity and non-commutativity in gene expression

In this section, we build models that provide mechanistic explanations for observations **(1, 2, 3, 4)**, emphasizing the non-additive and non-commutativity properties of two mutations on gene expression.

### 4.1 Mathematical background

First, consider ordinary differential equation (ODE) models for gene expression and regulation. For gene X with expression level *x*(*t*), the simplest model d*x/*d*t* = *λ* – *γx* considers only synthesis and degradation with constant rates *λ, γ*, and a stationary state *x*^∗^ = *λ/γ*. If other genes (mutations) regulate the expression of X, we can allow the synthesis rate *λ* to depend on other factors, which may include the activation state of genes Y and Z. For example, we might write a deterministic model for the expression level *x*(*t*) as

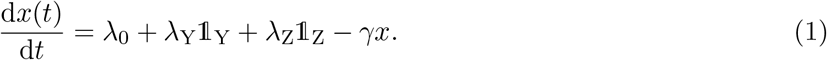

Here, we have modeled the synthesis rate *λ* = *λ*_0_ + *λ*_Y_𝟙_Y_ + *λ*_Z_𝟙_Z_ as a Boolean control operator with 𝟙_Y_ = 1 if Y (gene activity or product) is present, 𝟙_Y_ = 0 otherwise, and *λ*_Y_ is a constant regulation amplitude of gene Y on the expression of gene X. A similar term with amplitude *λ*_Z_ arises for mutation Z. After 𝟙_Y_ or 𝟙_Z_ changes (e.g., one gene mutates), the expression level of X will eventually return to a new equilibrium. Therefore, in this section, we only consider the stationary state *x*^∗^.

The linear (in *x*) ODE in Eq. 1 cannot explain observations **(1-4)** since the regulation effects of different genes (mutations) are additive and commutative. Regardless of the status of other genes and mutations, the presence of one mutation always has the same effect. Therefore, one needs to include a nonlinear term. Consider

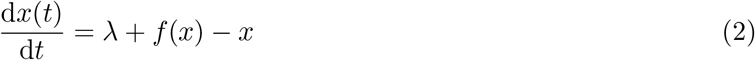

where for simplicity we have normalized time so that the intrinsic degradation rate *γ* ≡ 1 and *λ* is the dimensionless synthesis rate that may still depend on the presence of mutations of other genes (thus being externally tunable). The nonlinear term *f* (*x*) represents the autoregulation of *X* [55]. A possible form of *f* (*x*) is

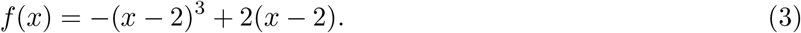

While many possible forms for *f* (*x*) may be inferred from measurements or otherwise approximated or modeled, we will use the form in Eq. 3 to explicitly illustrate the effects of such an autoregulation term on the expression of X. The fixed points (stationary states) of Eq. 2 using *f* (*x*) given in Eq. 3 are plotted in Fig. 1 as a function of *λ* and show the high and low expression level branches.

**Figure 1:**
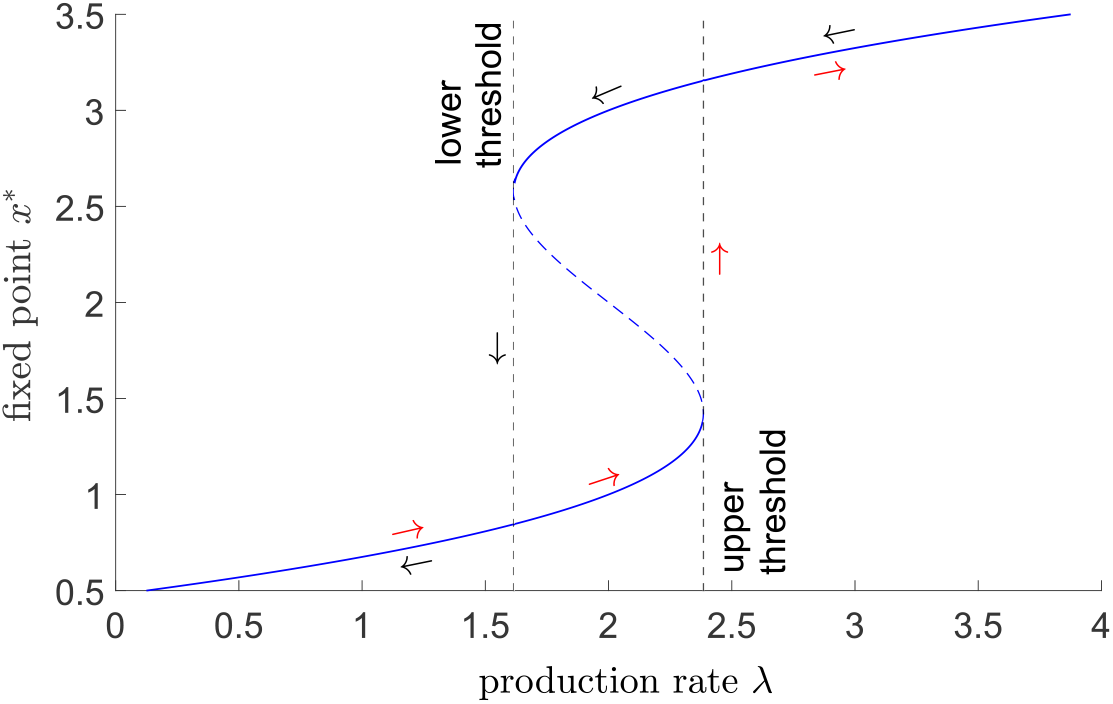
The fixed points of Eq. 2 for different values of the expression rate of gene X, *λ*. The solid blue line is the stable fixed point, and the dashed blue line is the unstable fixed point. If the system starts at *λ <* 1.6, the only fixed point is *x*^∗^ ≈ 0.8. As *λ* is increased, the system moves along the red arrow in the low branch until reaching the upper threshold at *λ* ≈ 2.4, at which point the system jumps to the high-*x*^∗^ branch, and the fixed point jumps from *x*^∗^ ≈ 1.4 to *x*^∗^ ≈ 3.2. If the synthesis rate is lowered starting from a value *λ >* 2.4, the system moves along the black arrow along the high-value branch until reaching the lower threshold at *λ* ≈ 1.6, at which the system jumps from *x*^∗^ ≈ 2.6 to the low-value branch at *x*^∗^ ≈ 0.8.

For this nondimensionalized model, when *λ <* 1.6, there is one stable, low-value fixed point at *x*^∗^ ≲ 0.8. If *λ >* 2.4, there is one stable fixed point *x*^∗^ ≳ 3.2 continued from the stable high-value branch. At intermediate values 1.6 *< λ <* 2.4, both values of *x*^∗^ (high and low) are locally stable and are connected by an unstable middle branch of fixed points (dashed curve).

When we start from *λ <* 1.6, the system resides only on the low expression level branch. If *λ* is then increased to 1.6 *< λ <* 2.4, although there are two stable branches, the system stays at the low-*x*^∗^ branch. When we further increase *λ* until *λ >* 2.4, the stable low-*x*^∗^ branch and the unstable middle branch collide and disappear (saddle-node bifurcation), and the system jumps to the stable high-*x*^∗^ branch. If we start with *λ >* 2.4, the system is at the high-level branch. Decreasing *λ* to 1.6 *< λ <* 2.4, the system will stay at the stable high level branch until *λ <* 1.6, when the stable high-level branch and the unstable intermediate-value branch collide and disappear, and the system jumps to the low-*x*^∗^ branch. In this model, when we change the parameter *λ* along different trajectories, even though they all arrive at final values 1.6 *< λ <* 2.4, the stationary state can differ. For example, if the value of *λ* is evolved according to *λ* = 2 → *λ* = 1 → *λ* = 2, the final state is *x*^∗^ = 1, but if *λ* follows the trajectory *λ* = 2 → *λ* = 3 → *λ* = 2, the final state is the high-value one at *x*_∗_ = 3.

Now consider a model in which the source of *X* is controlled by genes Y and Z through *λ* = *λ*_0_ + *λ*_Y_𝟙_Y_ + *λ*_Z_𝟙_Z_. Genes Y and Z can qualitatively affect the stationary state values of expression of X, *x*^∗^, if including their presence (or absence) induces *λ* to cross the thresholds at 1.6 and 2.4. This model structure means that different orders of mutations (changes in Y and Z) can give rise to different stationary states and lead to non-additive and non-commutative effects on X. We now use the model structure given by Eq. 2 to explain observations **(1, 2, 3, 4)**. Note that we just need Eq. 2 to be nonlinear (to generate non-additivity) and exhibit bistability (to induce non-commutativity).

### 4.2 Models for observations (1, 2)

We consider different variants of Eq. 2 to explain why some genes have 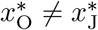, but 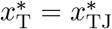 (and vice versa). In the following, “J” will indicate the JAK2 mutation while “T” will denote the TET2 mutation. In this application, Y and Z are identified as target genes regulated by J and T. Thus, we can simplify the expression rate *λ* in Eq. 2 to, *e*.*g*., *λ* = 0.5 + 𝟙_J_ + 𝟙_T_. With no mutation, *λ* = *λ*_0_ = 0.5, and the system is in the low-expression state 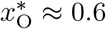. Consider a scenario in which 𝟙_T_ = 0, 𝟙_J_ = 1, *i*.*e*., the JAK2 mutation is present but not the TET2 mutation (or vice versa). Then, *λ* = 1.5 and the system is at the low-*x*^∗^ state 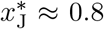 (also, 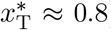). If both JAK2 and TET2 mutations are present, then 𝟙_T_ = 𝟙_J_ = 1, *λ* = 2.5, and the system is in the high-*x*^∗^ state 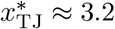. We have 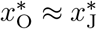 but 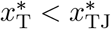. Therefore, in this case, JAK2 mutation up-regulates X only if the TET2 mutation is present. See Fig. 2(a) for an illustration of this scenario.

**Figure 2:**
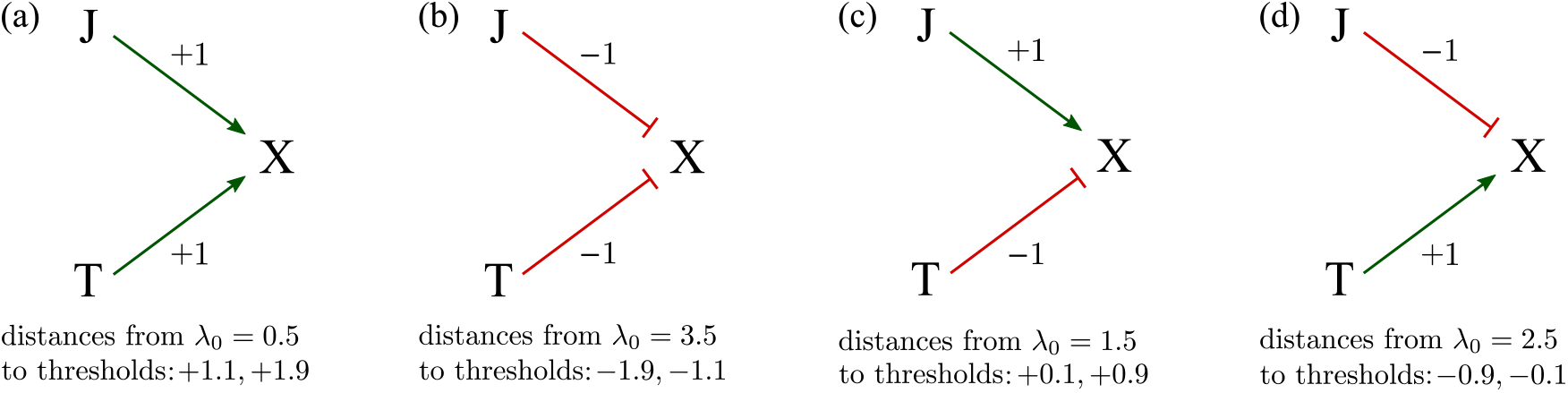
(a) A schematic of the model *λ* = 0.5 + 𝟙_J_ + 𝟙_T_ that yields 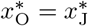 and 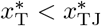. “J → X” indicates that the presence of a J mutation up-regulates expression of X. In this particular model, we have 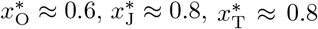, and 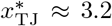. (b) Schematic of the model *λ* = 3.5 – 𝟙_J_ – 𝟙_T_ which yields 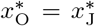 but 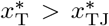. “J,T ⊣ X” indicates that JAK2 and TET2 mutations both down-regulate X. Here, we have 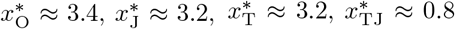. (c) The production rate model *λ* = 1.5 + 𝟙_J_ – 𝟙_T_ captures 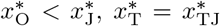. Here, we have 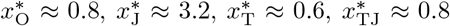. (d) *λ* = 2.5 – 𝟙_J_ + 𝟙_T_ explains 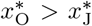 but maintains 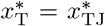. As before, the symbols “⟶” and “**⊣**” represent up-regulation and down-regulation, respectively. This scenario yields 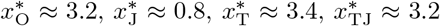. The distances of the lower and upper thresholds to the value of *λ*_0_ are indicated for all cases.

Now, assume the regulated production rate takes the form *λ* = 3.5 – 𝟙_J_ – 𝟙_T_. If 𝟙_T_ = 0 (no TET2 mutation), then *λ* = 3.5 and a JAK2 mutation itself does not affect X expression much (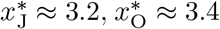). If 𝟙_T_ = 1, then a JAK2 mutation (changing 𝟙_J_ from 0 to 1) will alter the *x*-production rate to *λ* = 1.5, sufficient to decrease the steady state expression from 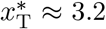 to 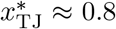. While 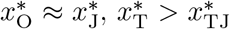. Thus, the JAK2 mutation down-regulates expression of X only if the TET2 mutation is present. See Fig. 2(b) for a schematic of this scenario.

Now, if *λ* = 1.5 + 𝟙_J_ – 𝟙_T_, then if the T is absent (no TET2 mutation), the presence of J (a JAK2 mutation) up-regulates X since 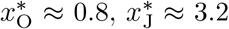. In the presence of T, J does not affect X expression much since 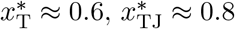. This regulation model is depicted in Fig. 2(c).

Finally, consider a gene expression rate governed by *λ* = 2.5 – 𝟙_J_ + 𝟙_T_, as shown in Fig. 2(d). If T is not present, then J down-regulates X since 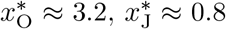. In the presence of T, J does not affect X expression much since 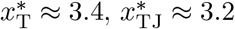.

### 4.3 Model for observation (3)

To explain observation **(3)** that O → J and T → TJ have opposite effects, we need a more complicated variant of Eq. 2. Consider a gene Y whose expression level *y* is described by

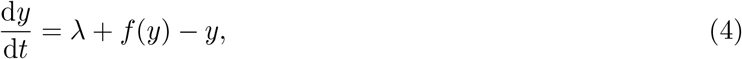

in which *λ* = 1.5 + 𝟙_J_ – 𝟙_T_ and *f* (*y*) = –(*y* – 2)^3^ + 2(*y* – 2). This setup gives rise to 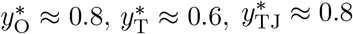, and 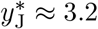. Now, consider a gene X whose expression level follows the linear dynamics

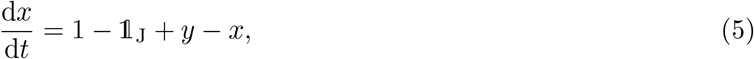

where X has a basal synthesis and decay rate of 1. A JAK2 mutation can directly down-regulate X expression with strength 1, while expression of Y can up-regulate that of X with strength proportional to its expression level *y*. Fig. 3(a) shows the key regulation processes in this model. Without JAK2 and TET2 mutations, *λ* = 1.5, which is under the lower threshold of *λ* = 1.6. In this case, Y is in its low-expression state 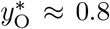 and X is only weakly affected by Y, with a stationary expression level 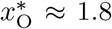. With J but not T, *λ* = 2.5, which is above the upper threshold 2.4. In this case, Y is in its high-value state 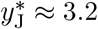. Now, X expression is affected by both J and Y (strongly), taking on the value 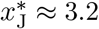. With T but not J, *λ* = 0.5, below the lower threshold of 1.6. In this case, Y is in its low-value state 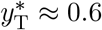 and X expression, 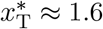, is only weakly affected by Y expression.

**Figure 3:**
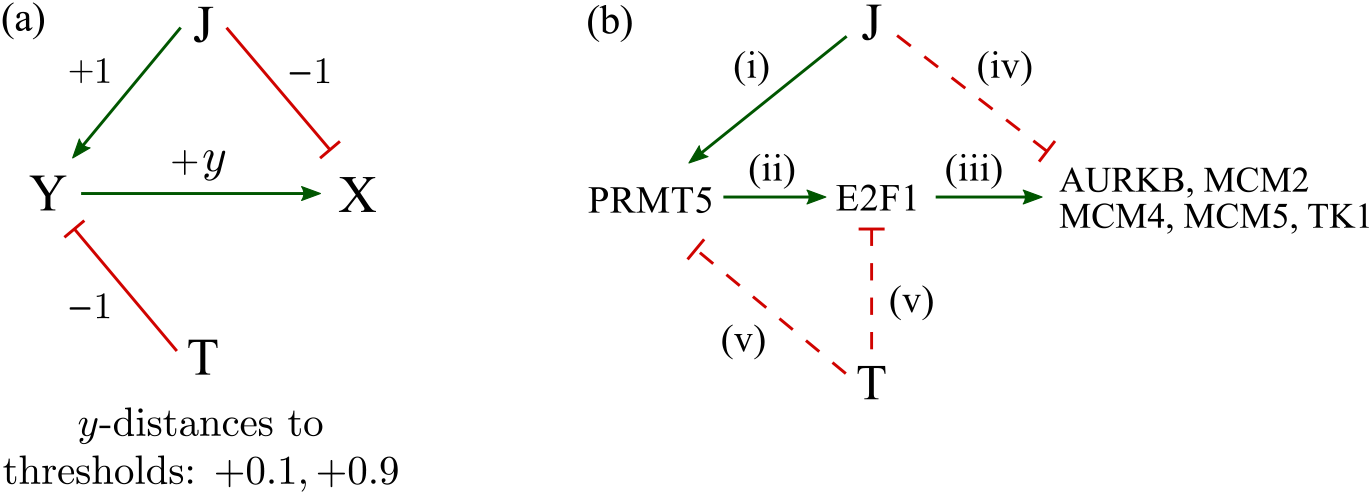
(a) Schematic of a model that explains 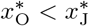 but 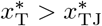. In this model, the steady-state expression levels of Y are 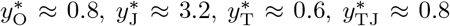. The basal value of *x* = 1, while the different stationary expression levels of X are 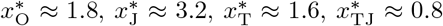. (b) The gene regulatory network that explains 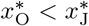 but 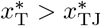 for a number of genes. Solid line indicates a verified regulation while the dashed line denotes a hypothesized regulatory interaction.

In the presence of both JAK2 and TET2 mutations, *λ* = 1.5, under the lower threshold of 1.6. In this case, Y is in its low-value state 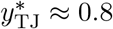 and X is affected weakly by Y expression and by the JAK2 mutation, with 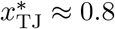. Therefore, without a TET2 mutation, JAK2 mutation up-regulates X expression (from 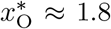 to 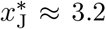); with the TET2 mutation, a JAK2 mutation down-regulates X expression from 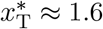 to 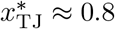.

This proposed model introduces an extra gene Y in order to explain 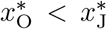 and 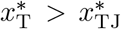. To connect our ODE model to observations of specific genes, and to find potential candidates for Y, we list some specific experimental findings:

i. For MPN, the expression of PRMT5 is increased in cells with the JAK2 V617F mutation [41].
ii. PRMT5 inhibition reduces the expression of E2F1. Thus, PRMT5 up-regulates E2F1 [41].
iii. The expression of E2F1 induces all genes of the endogenous MCM family [39]. E2F1 is a transcriptional activator of AURKB [58] that can up-regulate AURKB and MCM5 expression [45]. Overexpressing E2F1 alone results in the up-regulation of MCM5 and TK1 [28]. In sum, E2F1 up-regulates AURKB, MCM2, MCM4, MCM5, and TK1.
iv. From observations (a)-(c), JAK2 mutation indirectly up-regulates AURKB, MCM2, MCM4, MCM5, and TK1 through PRMT5 and E2F1. We propose that JAK2 mutation can weakly but directly down-regulate these genes. This hypothesis can be verified experimentally by introducing the JAK2 mutation after the knockdown or knockout of PRMT5 or E2F1 and observing a decreased expression of AURKB, MCM2, MCM4, MCM5, and TK1.
v. We propose that a mutated TET2 can down-regulate E2F1 directly, or indirectly through PRMT5. This down-regulation cancels out the up-regulation JAK2 → PRMT5 → E2F1. This means E2F1 (and possibly PRMT5) expression satisfies 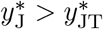 and 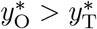.

In summary, in patients without a TET2 mutation, the JAK2 mutation can up-regulate PRMT5 and E2F1, which in turn up-regulate AURKB, MCM2, MCM4, MCM5, and TK1; this strong indirect upregulation of JAK2 → PRMT5 → E2F1 → AURKB/MCM2/MCM4/MCM5/TK1 can cover the weak direct down-regulation JAK2 ⊣ AURKB/MCM2/MCM4/MCM5/TK1, and the overall effect is 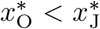. In the presence of the TET2 mutation, the up-regulation JAK2 → PRMT5 → E2F1 is covered by the down-regulation TET2 ⊣ PRMT5/E2F1; therefore, PRMT5 and E2F1 are locked to low levels so that the only effective regulation of JAK2 is the down-regulation JAK2 ⊣ AURKB/MCM2/MCM4/MCM5/TK1. This means we have 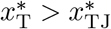.

Ortmann et al. [40] reported ten genes that follow 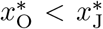 but also 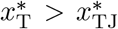: AURKB, FHOD1, HTRA2, IDH2, MCM2, MCM4, MCM5, TK1, UQCRC1, and WDR34. Our model can explain five of them (AURKB, MCM2, MCM4, MCM5, TK1) with the same pathway JAK2 → PRMT5 → E2F1 → AURKB/MCM2/MCM4/MCM5/TK1, while the role of Y can be played by E2F1 and/or PRMT5. Fig. 3(b) shows a simple gene regulatory network that is consistent with the observations. Our proposed model also implies a number of predictions. Specifically, the JAK2 mutation weakly but directly down-regulates genes that satisfy 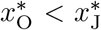 and 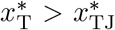. Besides, E2F1 and possibly PRMT5 have 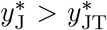 and 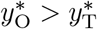.

The pathway JAK2 → PRMT5 → E2F1 → … is but one possibility. There is also evidence for the role of p53 in observation **(3)**. JAK2 V617F negatively regulates p53 stabilization [35], while p53 can regulate AURKB and MCM5 [45]. The complete gene regulatory network should be determined using certain inference methods based on gene expression data [57, 6].

### 4.4 Model for observation (4)

To explain observation **(4)** that TJ and JT have different effects, namely 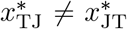, consider Eq. 2 with *λ* = 2 + 𝟙_J_ – 𝟙_T_. With J but not T, *λ* = 3 and X lies in its only high-value stationary state 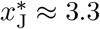 if T appears after J, then *λ* = 2, and X remains in its high-value branch with stationary level 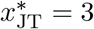. If the TET2 mutation arises with a JAK2 mutation, *λ* = 1 and the steady-state expression of X is 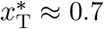; if J appears after T, then *λ* = 2 and X expression remains in its low-value branch with stationary value 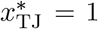. See Fig. 1 for a more detailed description. For MPN patients, if the order is JT, the final X expression is high (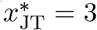); if the order is TJ, the final X expression level is low (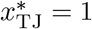). See Fig. 4(a) for an illustration of this model explaining 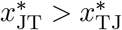.

**Figure 4:**
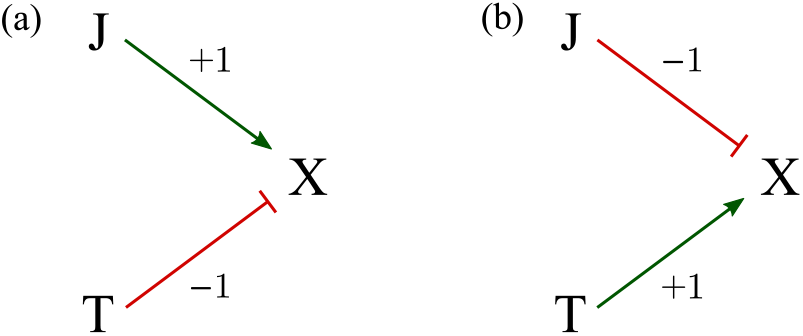
(a) A schematic of the model *λ* = 2 + 𝟙_J_ – 𝟙_T_ in Eq. 2 which explains 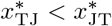. If the effects of JAK2 and TET2 mutations towards the input *λ* are together greater than 0.4 (*i*.*e*., with JAK2 but not TET2), the system is forced to be on the high-*x*^∗^ branch; if the contribution to *λ* input JAK2 and TET2 is smaller than –0.4 (*i*.*e*., with TET2 but not JAK2), the system ends up on the low-value branch. (b) The model *λ* = 2 – 𝟙_J_ + 𝟙_T_ can yield 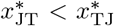. If the contribution from JAK2 and TET2 mutations to *λ* is greater than 0.4 (i.e., with TET2 but not JAK2), the system is forced onto the high-*x*^∗^ branch; if the JAK2 and TET2 contributions to the input *λ* is smaller than –0.4 (i.e., with JAK2 but not TET2), the system is forced onto the low-*x*^∗^ branch.

To explain 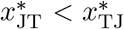, consider Eq. 2 with *λ* = 2 – 𝟙_J_ + 𝟙_T_. If the mutation order is JT, the final X expression level is low (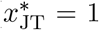); if the order is TJ, the final X expression level is high (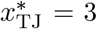). This regulation control mechanism is illustrated in Fig. 4(b).

## 5 Models for non-commutativity in cell population and age

In this section, we build models to explain observations **(5, 6)** that the age of diagnosis and the populations of cancer cells depend on the order of the two mutations experienced by the patient. Clinically, the mutations are not commutative.

### 5.1 Different mechanisms for explaining observations (5, 6)

For observations **(5, 6)**, the cell population and age are measured at the time of diagnosis. However, it is difficult to know the time interval between acquiring the second mutation and diagnosis or to model the disease progression during this time. Therefore, we analyze observations **(5, 6)** focussing on the time the first double-mutation cell (with both JAK2 and TET2 mutations) appears. Doing so, we must assume an amendment to the observations:

**(5’)** For TET2-first patients, at the time when the first TET2-JAK2 cell appears, the percentage of TET2-only cells is significantly higher than the percentage of JAK2-only cells at the time when the first JAK2-TET2 cell appears in JAK2-first patients.

**(6’)** For JAK2-first patients, the time at which the first JAK2-TET2 cell appears is significantly earlier than the time at which the first TET2-JAK2 cell appears for TET2-first patients.

Thus, we are assuming that the time delay between the appearance of the first double-mutation cell and diagnosis is independent of the order of mutation. In this case, observation **(6)** and observation **(6’)** are equivalent.

The relationship between observation **(5)** and observation **(5’)** is complicated. After the appearance of one mutation, gene expression levels can reach the new stationary states relatively quickly, typically within a cell lifespan. However, *populations of cells* with different mutations may take years before reaching steady-state numbers (*e*.*g*., for cells with both JAK2 and TET2 mutations to dominate). Therefore, between the appearance of the first double-mutation cell and diagnosis, the cell population composition may have changed significantly. Nevertheless, we assume that the percentage of JAK2-only cells in JAK2-first patients and the percentage of TET2-only cells in TET2-first patients does not change appreciably before diagnosis. In this sense, observations **(5)** and **(5’)** can be assumed equivalent.

A. Ortmann et al. [40] propose that *cells with a JAK2 mutation have only a mild proliferation advantage while cells with a TET2 mutation (whether JAK2 is present or not) have a more significant proliferation advantage*. This feature would explain observation (**5’**). If the JAK2 mutation first appears, such JAK2-only cells proliferate only slightly faster than non-mutant cells. Thus, there are few JAK2-only cells that can acquire the TET2 mutation. If the TET2 mutation appears first, such TET2-only cells grow much faster than non-mutant cells. Thus, there is a higher population of TET2-only cells when the JAK2 mutation appears. In the following simulations, we find that this mechanism can also be used to explain observation (**6’**). The model by Teimouri and Kolomeisky [52] is relevant to this mechanism in that they assume different proliferation rates between JAK2-only mutated cells and TET2-only mutated cells, but assume equal proliferations rates for JT and TJ cells. They incorporate a number of assumptions that are not satisfied in this system.
B. Since different mutations generally appear with different rates [32], a more general model can also include different rates for the different mutations. Here, we propose a mechanism in which *JAK2 and TET2 have different mutation rates*, explaining observations **(5’)** and **(6’)**. If the mutation rate of JAK2 is lower than that of TET2, then when JAK2 appears first, JAK2-only cells have a shorter time to proliferate before a TET2 mutation appears. This provides a basis for observation **(5’)**. The explanation for observation **(6’)** is given in Subsection 5.4 below.
C. We propose a cooperative mutation mechanism that can also lead to **(5’)** and **(6’)**: cells with the JAK2 mutation carry a higher mutation rate for TET2 mutation. In other words, *a JAK2 mutation can induce an additional TET2 mutation*. Therefore, a TET2 mutation can arise quickly after the first JAK2 mutation appears, explaining observation **(6’)**. JAK2-only cells do not have much time to proliferate (into JAK2-only daughter cells) before the appearance of a TET2 mutation, consistent with observation **(5’)**.

### 5.2 Generalized Moran process

We implement a simple Moran population model to explore consequences of mechanisms **(A), (B)**, and **(C)**. Cell population dynamics that include state transitions have been widely studied [59, 38, 8, 2]. To mathematically model observations **(5’)** and **(6’)**, we consider a simple discrete-time Moran model [16, 43], shown in Fig. 5, for cell populations that include mutations. A continuous-time Moran model can also be straightforwardly constructed and analyzed. A related two-mutation branching process has formulated describe the first times to acquire double mutations, but did not distinguish the order of mutation acquisition [11]. Moreover, unlike branching processes [23], the total number of cells is fixed in our Moran process. This is a reasonable approximation for stable hematopoietic stem cell populations and allow us to easily estimate relative populations of all cells. We will assume that cells can exist in five states: non-mutant, JAK2-only, TET2-only, JAK2-TET2, and TET2-JAK2. Here, cells with two mutations, for example JAK2-TET2, are those that are part of a lineage that was started when a single JAK2-mutation mother cell divided into daughter cells that acquired the TET2 mutation.

**Figure 5:**
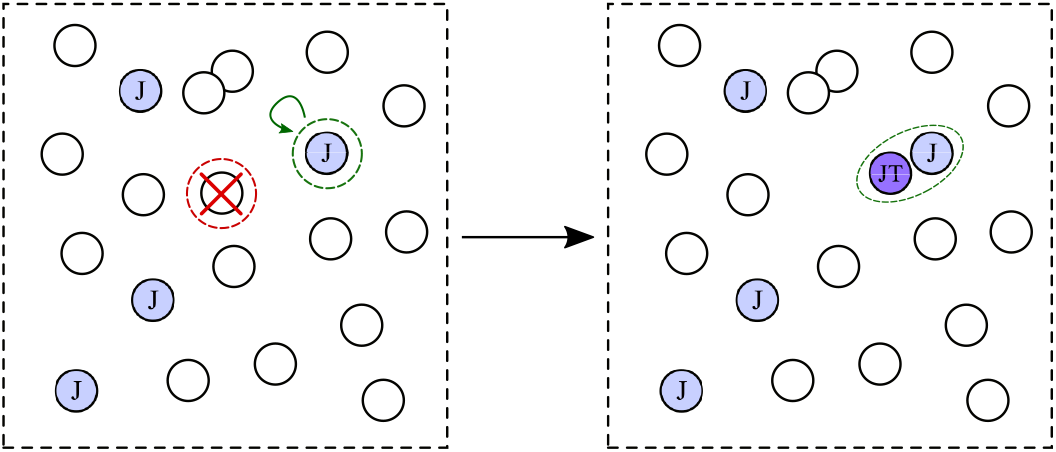
A schematic of the steps in our Moran process. At some time, the system contains sixteen wild-type cells and four JAK2-mutated cells. In each timestep, one cell (wild-type) is chosen for removal (red-dashed circle), while another (J) is chosen for replication (green-dashed circle), during which one daughter may acquire a mutation. In this example, a J cell divides into a J cell and a double-mutant JT cell, thus defining the end point of our simulation.

In the following, the suffix O denotes wild-type cells, J denotes JAK2-only cells, T describes TET2-only cells, JT defines JAK2-TET2 cells, and TJ labels TET2-JAK2 cells. The number of wild-type cells is *n*_O_ and the relative birth and death coefficients of these unmutated cells are *b*_O_ and *d*_O_, respectively. The analogous populations and birth and death coefficients are similarly defined for J, T, JT, TJ-type cells. At each time step, one cell is randomly–weighted by the death rate of its type–picked for removal. Simultaneously, another cell is randomly–weighted by its birth rate–picked for replication. After division, one daughter cell will remain in the same state as the mother cell, while the other may transform into another type according the corresponding mutation probability. There are four possible mutation probabilities *m*_O→J_, *m*_O→T_, *m*_J→JT_, and *m*_T→TJ_. For example, *m*_O→J_ is the probability that the chosen daughter cell of a wild-type mother cell acquires the JAK2 mutation. The state space of this process is (*n*_O_, *n*_J_, *n*_T_, *n*_JT_, *n*_TJ_), with a total fixed-population constraint Σ_*j*_ *n*_*j*_ = *n*.

At each time point, the probability that a wild-type cell is chosen for elimination is

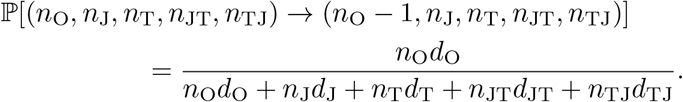

The probability of selecting other cell types for death are similarly defined. The probability that a wild-type cell is chosen to divide, and that no mutations arise in the daughter cells is

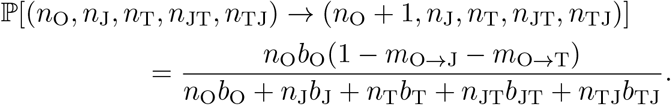

Similarly, the probabilities of generating additional cells of other cell types are

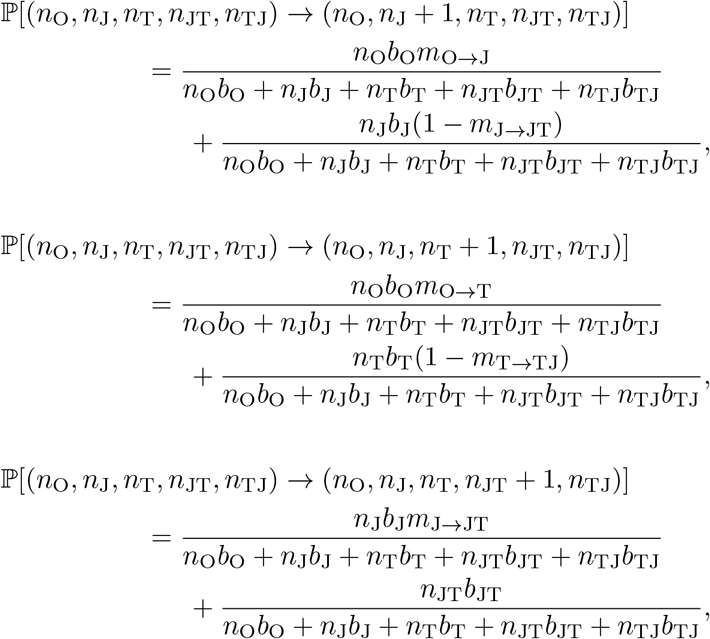

and

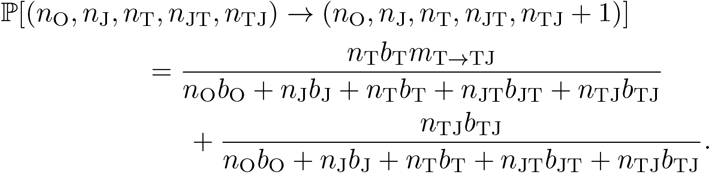

### 5.3 Simulation results

In the simulations, for all three mechanisms, we set the initial population *n*_O_ = 100, *n*_J_ = *n*_T_ = *n*_JT_ = *n*_TJ_ = 0, so that the total population is *n* = 100. We set a common value for all death probabilities *d*_O_ = *d*_J_ = *d*_T_ = *d*_JT_ = *d*_TJ_ = 1. Table 3 lists the relative birth rates and mutation probabilities associated with three different mechanisms.

**Table 3:**
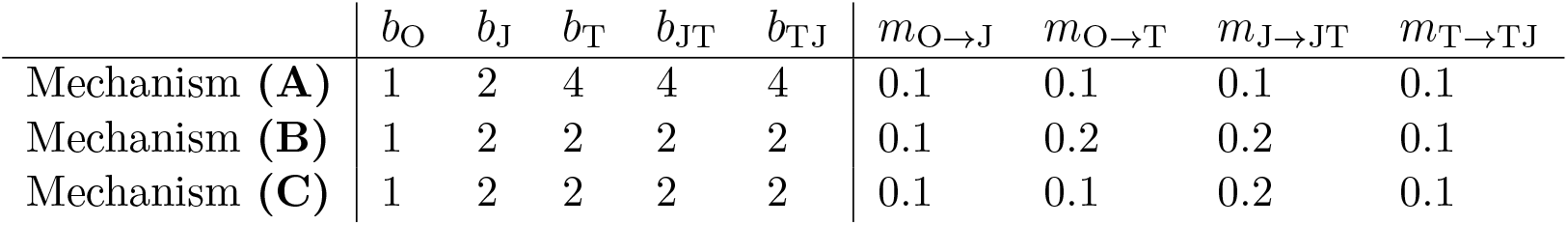
A generalized Moran process is simulated to explore three different mechanisms, **(A), (B)**, and **(C)**, with corresponding relative birth coefficients *b* and mutation probabilities *m* listed. The mutation probability is the probability that one daughter acquires a mutation at birth.

| | $b_O$ | $b_J$ | $b_T$ | $b_{JT}$ | $b_{TJ}$ | $m_{O \rightarrow J}$ | $m_{O \rightarrow T}$ | $m_{J \rightarrow JT}$ | $m_{T \rightarrow TJ}$ |
| --- | --- | --- | --- | --- | --- | --- | --- | --- | --- |
| Mechanism **(A)** | 1 | 2 | 4 | 4 | 4 | 0.1 | 0.1 | 0.1 | 0.1 |
| Mechanism **(B)** | 1 | 2 | 2 | 2 | 2 | 0.1 | 0.2 | 0.2 | 0.1 |
| Mechanism **(C)** | 1 | 2 | 2 | 2 | 2 | 0.1 | 0.1 | 0.2 | 0.1 |

In Mechanism **(A)**, all mutations have the same probability and cells with the TET2 mutation proliferate faster than those with JAK2-only mutations. In Mechanism **(B)**, all cells with at least one mutation have the same relative birth coefficients, while the appearance probability of a TET2 mutation is set higher than that of a JAK2 mutation. In Mechanism **(C)**, all cells with at least one mutation have the same relative birth coefficients, but a cell with a JAK2 mutation more likely acquires a TET2 mutation upon division, corresponding to a larger *m*_J*→*JT_.

Since we use this Moran process to study observations (**5’, 6’**) at the time that the first double-mutation cell appears, the process is stopped once *n*_JT_ = 1 or *n*_TJ_ = 1. At this point, if both *n*_J_ *>* 0 and *n*_T_ *>* 0 the order of mutation cannot be inferred and this simulation result is abandoned. If *n*_JT_ = 1 and *n*_T_ = 0, we record the corresponding *n*_J_ and the current time point *T*. This mechanism reflects a JAK2-first patient. If *n*_TJ_ = 1 and *n*_J_ = 0, we record the corresponding *n*_T_ and the current time point *T*. This mechanism reflects a TET2-first patient. For each mechanism, 10^6^ trajectories were simulated. Although most trajectories are abandoned, at least 10^4^ trajectories remained from which JAK2-first and TET2-first dynamics could be determined and sufficient statistics extracted.

To verify observation (**5’**), we compare 𝔼 (*n*_J_ | *n*_JT_ = 1, *n*_T_ = 0) and 𝔼 (*n*_T_ | *n*_TJ_ = 1, *n*_J_ = 0). To investigate observation (**6’**), we compare 𝔼 (*T* | *n*_JT_ = 1, *n*_T_ = 0) to 𝔼 (*T* | *n*_TJ_ = 1, *n*_J_ = 0). We also use a *t*-test to examine whether the difference in mean cell populations are significant. For each scenario, we run the simulation 10^6^ times.

For Mechanism **(A)**, 𝔼 (*n*_J_ | *n*_JT_ = 1, *n*_T_ = 0) = 4.02 *<* 5.41 = 𝔼 (*n*_T_ | *n*_TJ_ = 1, *n*_J_ = 0), and the *p*-value from the *t*-test is smaller than 10^*−*200^. We also find 𝔼 (*T* | *n*_JT_ = 1, *n*_T_ = 0) = 24.15 *<* 25.93 = 𝔼 (*T* | *n*_TJ_ = 1, *n*_J_ = 0), with a *t*-test *p*-value *∼* 10^*−*40^. This scenario can generate observation (**6’**). Since *b*_T_ *> b*_J_, the probability that *n*_JT_ = 1, *n*_T_ = 0 is smaller than that of *n*_TJ_ = 1, *n*_J_ = 0. Simulations with *n*_JT_ = 1, *n*_T_ = 0 generally mean that the JAK2 mutation happens to arise more quickly, and that these JAK2-only cells happen to divide more frequently. Under Mechanism **(B)**, 𝔼 (*n*_J_ | *n*_JT_ = 1, *n*_T_ = 0) = 2.37 *<* 6.51 = E(*n*_T_ | *n*_TJ_ = 1, *n*_J_ = 0) and 𝔼 (*T* | *n*_JT_ = 1, *n*_T_ = 0) = 11.52 *<* 24.75 = 𝔼 (*T* | *n*_TJ_ = 1, *n*_J_ = 0), both with a *t*-test *p*-value less 10^*−*200^. Finally, in Mechanism **(C)**, 𝔼 (*n*_J_ | *n*_JT_ = 1, *n*_T_ = 0) = 3.59 *<* 4.27 = 𝔼 (*n*_T_ | *n*_TJ_ = 1, *n*_J_ = 0) with *p*-value ~ 10^*−*192^ and 𝔼 (*T n*_JT_ = 1, *n*_T_ = 0) = 22.79 *<* 25.97 = 𝔼 (*T n*_TJ_ = 1, *n*_J_ = 0) with *p*-value ~ 10^*−*133^. After applying the Bonferroni correction to these six tests, we find the probability of rejecting at least one of the above results is *α* = 10^*−*39^.

In this model, we see that all three mechanisms can produce observations (**5’, 6’**). Biologically, it is natural to assume that JAK2 and TET2 have different mutation rates (Mechanism **(B)**). Mechanisms **(A)** and **(C)** require more supporting evidence, so we primarily propose the mechanism associated with **(B)**, which is sufficient to explain observations.

### 5.4 Theoretical analysis of Mechanism (B) for observation (6’)

A generalized Moran process model is difficult to study analytically. To explain why Mechanism **(B)** produces observation (**6’**), we consider a simplified model, which is a limiting mechanism of the generalized Moran model.

Assume *b*_J_ = *b*_T_ = *b*_JT_ = *b*_TJ_ ≫ *b*_O_. This means that once one mutation appears, cells with this mutation will dominate the population. Thus, the situation in which both populations are appreciable, *n*_J_ *>* 0 and *n*_T_ *>* 0, does not arise. If we further assume *m*_O*→*J_ = *m*_T*→*TJ_ *≡ m*_1_, *m*_O*→*T_ = *m*_J*→*JT_ *≡ m*_2_ and that *m*_1_ *< m*_2_ ≪ 1, we can approximate the distribution of times (number of time steps in our discrete-time simulations) *T*_1_ for JAK2 mutation to appear by an exponential distribution with parameter *m*_1_ so that 𝔼 (*T*_1_) *≈* 1*/m*_1_. The time *T*_2_ for a TET2 mutation to appear is also exponentially distributed with parameter *m*_2_ so that 𝔼 (*T*_2_) *≈* 1*/m*_2_, and *T*_1_ and *T*_2_ are approximately independent. *T*_1_ *< T*_2_ corresponds to the JAK2-first scenario, while *T*_1_ *> T*_2_ results in a TET2-first observation. Now, assume a faster TET2 mutation rate (*m*_2_ *> m*_1_) and define *T* = max{*T*_1_, *T*_2_} as the time at which both mutations first arise. We find, approximating in continuous-time,

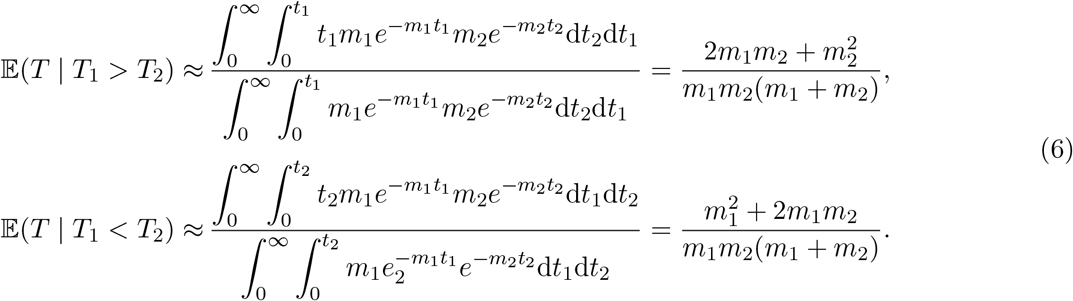

Since *m*_2_ *> m*_1_, 𝔼 (*T* | *T*_1_ *> T*_2_) *>* E(*T* | *T*_1_ *< T*_2_). For JAK2-first patients (*T*_1_ *< T*_2_), the waiting time for both mutations to appear is shorter than that for TET2-first patients (*T*_1_ *> T*_2_). One explanation for this is that when *T*_1_ *< T*_2_, it is more likely that *T*_1_ is exceptionally shorter, not that *T*_2_ is exceptionally longer.

## 6 Discussion and Conclusions

In this paper, we consider two genetic mutations in MPN: JAK2 and TET2. The effect of one mutation depends on whether the other mutation is present. When both the mutations are present, the order of their appearance also affects gene expression. For MPN, the order of the JAK2 V617F and DNMT3A mutations can also affect cellular proliferation [37]. The TET2 and DNMT3A mutations confer epigenetic changes in transcription that are passed on to daughter cells, thus providing a mechanism of “memory” required for bi/multistability and ultimately an order-of-mutation effect. Dependence of cell populations on the order of mutation also appear in other types of cancer. For example, in adrenocortical carcinomas, if the Ras mutation appears before the p53 mutation, the tumor will be malignant and metastatic, but if the p53 mutation appears before the Ras mutation, the tumor will be benign [21]. Similar observations can be found in other contexts [29, 53, 7].

We constructed several sub-models to explain the features of order-of-mutation effects in cancer, specifically addressing observations recorded to date for the JAK2/TET2 mutation pair in MPN. In Subsection 4.3, we describe experimental evidence that partially verifies our model. We also provided conjectures that can be tested experimentally: JAK2 mutation can weakly down-regulate AURKB, MCM2, MCM4, MCM5, and TK1 directly; TET2 mutation can down-regulate E2F1 and/or PRMT5.

Although we have developed a mathematical framework consistent with all observations to date, there are other possible processes that can lead to the rich set of observations discussed. Potential interactions with the adaptive immune system may inhibit cancer progression [34, 1]. Cancer may also inhibit the proliferation of white blood cells [19], which can lead to multistability in mathematical models of immune response to cancer [18, 31, 54]. Since certain mutations can help cancer cells escape the immune system [20], it is possible that the order of mutations affects cancer cell populations indirectly by interfering with the immune system. Finally, cancer cells can also affect and be affected by their microenvironments and other cells (through *e*.*g*., epigenetically driven “microenvironment feedback”). These nonlinear interactions have been modeled can lead to nonlinear dynamics in relative populations of different cancer cell types (different epigenetic or mutational states) [48]. Further developing models that incorporate immune and indirect cell-cell interactions could potentially lead to non-additivity and non-commutivity of mutation order in both gene expression and cell population dynamics. Formulating such mathematical frameworks, especially those coupling intracellular state dynamics to proliferating cell population will be the subject of future investigation.

## Data Availability

All data produced in the present study are available upon reasonable request to the authors.

## Acknowledgements

YW and TC acknowledge support from the National Institutes of Health through grant R01HL146552.

## A Detailed review of previously known models

Ortmann et al. [40] assume that TET2 mutation can significantly increase the proliferation rate of cancer stem cells, while JAK2 mutation only has a weak growth advantage. Therefore, for TET2-first patients, TET2-only cells first spread, and TET2-JAK2 cells (which do not have a significant growth advantage over TET2-only cells) do not dominate. For JAK2-first patients, JAK2-only cells do not spread that much, while JAK2-TET2 cells (after they appear) can dominate. Therefore, TET2-first patients have a much higher percentage of cells with only one mutation, consistent with observation **(5)**. In Section 5, we also discuss that these assumptions can explain observation **(6)**. See also the interpretation by Swanton [49].

Kent and Green [24] propose two explanations for observation **(4)**.

i. Both JAK2 and TET2 mutations can participate in epigenetic regulation [13, 22, 47], but the regulation mechanism might be incompatible. For example, the first mutation might lead to the occlusion of certain genomic regions, so that the second mutation cannot regulate genes in those regions. This mechanism would lead to 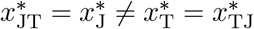, which is not consistent with other observations.
ii. For either JAK2-first or TET2-first patients, before the appearance of the second mutation, different first mutations might lead to different cell types and abundances, leading to different microenvironments in which double mutant cells that subsequently arise find themselves. This indirect effect can also shape disease progression.

In a related study, Roquet et al. [46] consider the effect of recombinases (i.e., genetic recombination enzymes) on gene sequences. When applying different recombinases to gene sequences, their order of application can lead to different results. For example, consider a gene sequence 12312 and two recombinases A, and B. Suppose A deletes genes between the “1s”, and B inverts genes between the two “2s” (if there are not two “2s”, B does nothing). If the DNA is exposed to A before B, the gene sequence becomes

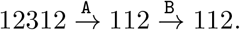

If A is added after B, the gene sequence becomes

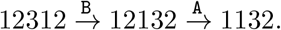

Although this system describes rearrangements and not specific mutations, it nonetheless provides a possible mechanism for observation **(4)**.

Ascolani and Liò [3] constructed a complex cellular automata model to describe cancer metastasis. They assume that different mutations lead to different proliferation and/or apoptosis rates. This assumption, as was invoked by Ortmann et al., can explain observation **(5)**. However, although Ascolani and Liò also take into account how different *orders* of different mutations can affect proliferation and apoptosis differently, they did not explicitly apply their model to the observations associated with different orders of JAK2 and TET2 mutations in the MPN system.

Clarke et al. [12] model the gene regulatory network as a generalized boolean network that evolves under certain rules and exhibits fixed points and/or limit cycles for gene expression levels. Each mutation fully activates or inhibits one gene (the expression level is fixed, similar to the do-operator used in causal inference [5]), thus changing the fixed points and/or limit cycles. Certain combinations of mutations lead to higher proliferation rates or apoptosis rates, making these mutation patterns more likely or less likely, respectively. There might be multiple fixed points and/or limit cycles, and different orders of mutations might lead to different final states of gene expression. Different sequences of perturbations leading to different states have been hypothesized for different physiological dynamics, including neuroendocrine stress response [26, 9]. This type of model can be used to explain observations **(4)** and **(5)**.

Talarmain et al. [51, 50] apply the model in Clarke et al.’s [12] paper to the JAK2/TET2 mutation order problem to explain observation **(4)**. They find a concrete generalized boolean network of gene expression. Mutations can affect the dynamics of this network. When there is no mutation or just one mutation, the system has one stable fixed point. When both JAK2 and TET2 mutations are present, the system has two stable fixed points. Different orders of mutations lead to different fixed points. The Talarmain et al. model invokes a third, downstream gene HOXA9 which is directly affected by different orders of JAK2 and TET2 mutations, which then affects many other downstream genes.

Mazaya et al. [33] use a boolean network to model the effect of mutations. The model dynamics and the explanation of observation **(4)** are similar to that of Clarke et al.’s model. Mazaya et al. further analyze this model to study when the network is more sensitive to different orders of mutations.

Finally, Teimouri and Kolomeisky [52] use a random walk model to study the acquisition of two mutations. The first stage is a random walk on 0, 1, …, *n*, representing the number of cells with the first mutation. The first stage starts at 0, and finishes when reaching *n*, meaning that all *n* cells have the first mutation. The process terminates if reaching 0 again before reaching *n*. The second stage is a random walk on *n, n* + 1, …, 2*n*, representing the number of cells with the second mutation plus *n*. The second stage starts at *n*, and finishes when reaching 2*n*, meaning that all cells have both mutations. If the process reaches 2*n*, we count the total time and take the expectation. They prove that if the first mutation has a higher fitness than the second mutation, then the tumor formation probability is higher, but the time for tumor formation is longer. This explains observation **(6)**.

### B Markov chain model for observation (4)

In the nonlinear ODE model for observation **(4)**, a cell with no mutation and a cell with both mutations carry the same gene expression landscape. Since gene expression at the single-cell level is essentially stochastic, we can also build an *ad hoc* Markov chain model in which the landscape of gene expression changes with the appearance of each mutation.

In a single cell, the expression level (protein or mRNA count) of gene X is a random variable *X* defined on 0, 1, 2, .... Define

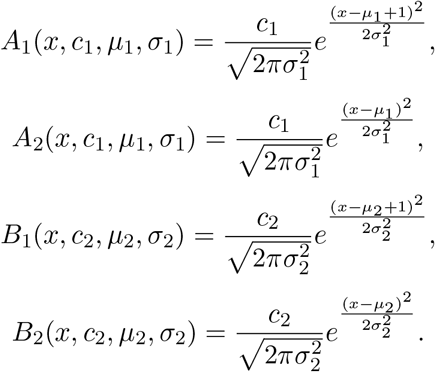

*X* follows a continuous-time Markov chain on 0, 1, 2,.... where the transition rate from *x* to *x* + 1 is

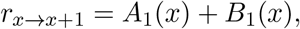

and the transition rate from *x* + 1 to *x* is

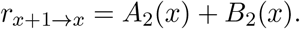

In all cases, we set *μ*_1_ = 1000, *μ*_2_ = 2000. If the JAK2 mutation is not present, we set *c*_1_ = 1 and *σ*_1_ = 400 in *A*_1_(*x, c*_1_, *μ*_1_, *σ*_1_) and *A*_2_(*x, c*_1_, *μ*_1_, *σ*_1_); otherwise, when the JAK2 mutation is present, we set *c*_1_ = 5 and *σ*_1_ = 80. Similarly, if TET2 mutation is not present, we set *c*_2_ = 1 and *σ*_2_ = 400 in *B*_1_(*x, c*_2_, *μ*_2_, *σ*_2_) and *B*_2_(*x, c*_2_, *μ*_2_, *σ*_2_); otherwise, when the TET2 mutation is present, we set *c*_2_ = 5 and *σ*_2_ = 80.

Since this Markov chain has no cycles, the detailed balance condition is satisfied [56], and we can directly compute the stationary probability distribution ℙ (*X* = *x*) from

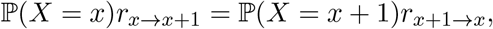

which is plotted in Fig. 6. If no mutation is present, the stationary distribution is rather flat with two low peaks near *x* = 1000 and *x* = 2000, as shown in (a). If only the JAK2 mutation is present, the stationary distribution is mostly concentrated in a sharp peak near *x* = 1000, with a small flat probability peak near *x* = 2000, as depicted in (b). If the TET2 mutation appears after the JAK2 mutation, the small flat probability peak near *x* = 2000 first sharpens to a more localized peak near *x* = 2000 (d); after an unrealistically long time (*e*.*g*., thousands of years), the heights of two peaks near *x* = 1000 and *x* = 2000 equalize, as shown in (f). If only the TET2 mutation is present, the stationary distribution shown in (c) is mostly concentrated in a sharp peak near *x* = 2000, with a small nearly flat probability mound near *x* = 1000. If the JAK2 mutation then appears, the small broad probability peak near *x* = 1000 first shrinks to a sharper peak near *x* = 1000 (e); after an extremely long time, the heights of two peaks near *x* = 1000 and *x* = 2000 again equilibrate as shown in (f).

**Figure 6:**
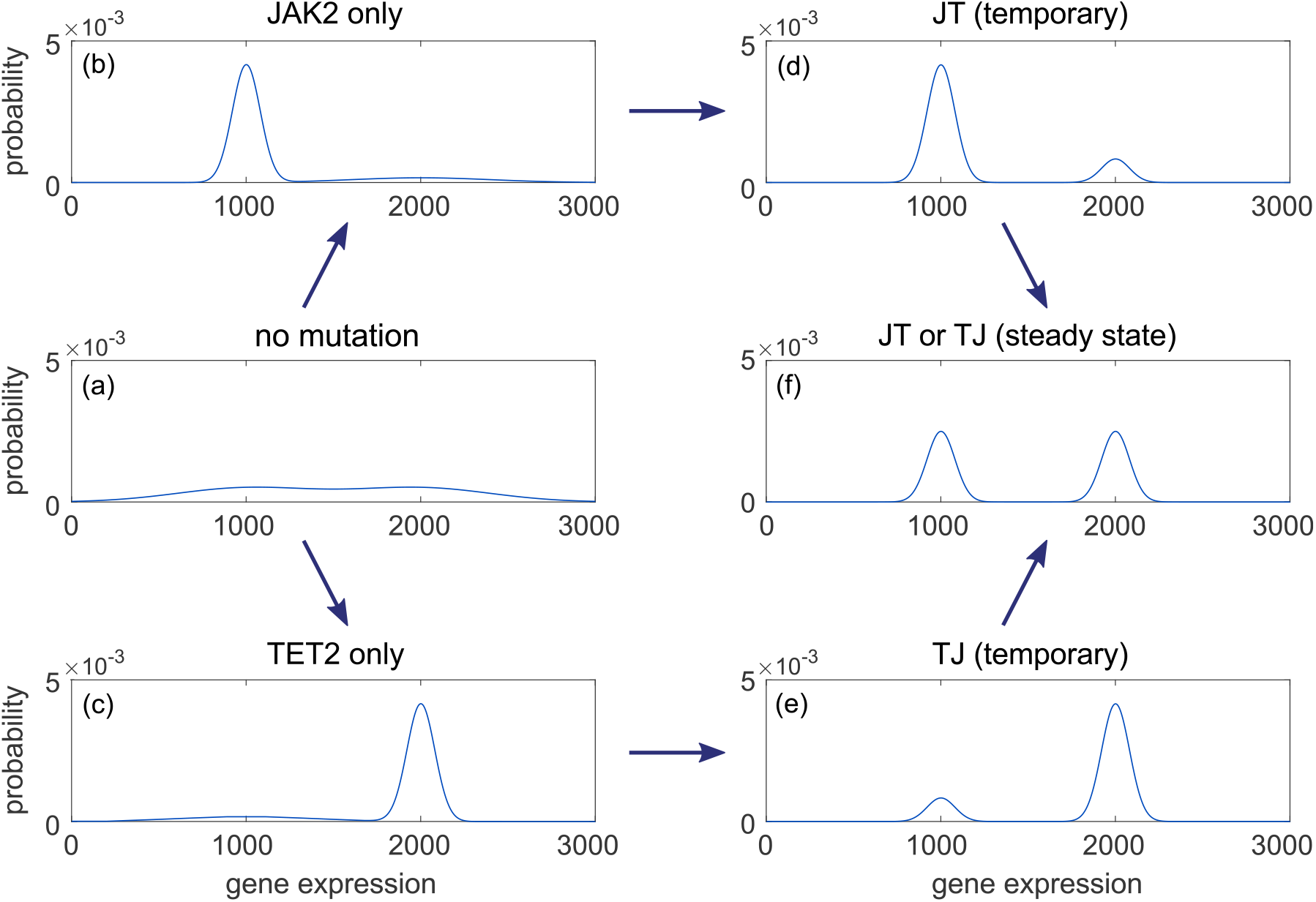
The probability distribution over gene expression levels *x* under the different enumerated scenarios described by the Markov chain model in Appendix B. (a) The stationary distribution when no mutation is present. (b) The distribution of *x* in cells with only a JAK2 mutation. (c) The stationary distribution when only TET2 mutation is present. (d) The temporary (but long-lived) distribution when a TET2 mutation appears on a lineage with an existing JAK2 mutation. (e) Long-lived probability distribution of *X* if the JAK2 mutation appears on a background of existing TET2 mutations. (f) The final distribution when both JAK2 and TET2 mutations are present (regardless of order).

We can also define a potential at *X* = *x* as the negative logarithm of the stationary distribution: *U* (*x*) = *−* log ℙ(*X* = *x*). Fig. 7 shows the potential function *U* (*x*) corresponding to different temporal configurations of mutations. If no mutation is present, the potential has two shallow wells near *x* = 1000 and *x* = 2000. The expression level can easily move between these two wells, as shown in Fig. 7(a). Fig. 7(b) depicts the case in which only the JAK2 mutation is present for which there is a deep well near *x* = 1000 and a shallow well near *x* = 2000. Here, it is easy to jump from the shallow well into the deep well, but not the other way around. Thus, this system is most likely to have expression level *x* = 1000. If the TET2 mutation appeared after the JAK2 mutation (Fig. 7(d)), the system will first stay in the deep well near *x* = 1000. Since both wells are deep, there is very little probability flux from one well to another and the probability distribution relaxes very slowly (over times longer than the life span of a human) towards final equipartition. If only the TET2 mutation is present, the system is likely to stay in the deep well near *x* = 2000, indicated in Fig. 7(c). Finally, if the JAK2 mutations appear after TET2 mutations, the system will reside in the well near *x* = 2000 (see Fig. 7(e)) before very slowly becoming equally distributed between *x ≈* 1000 and *x ≈* 2000.

**Figure 7:**
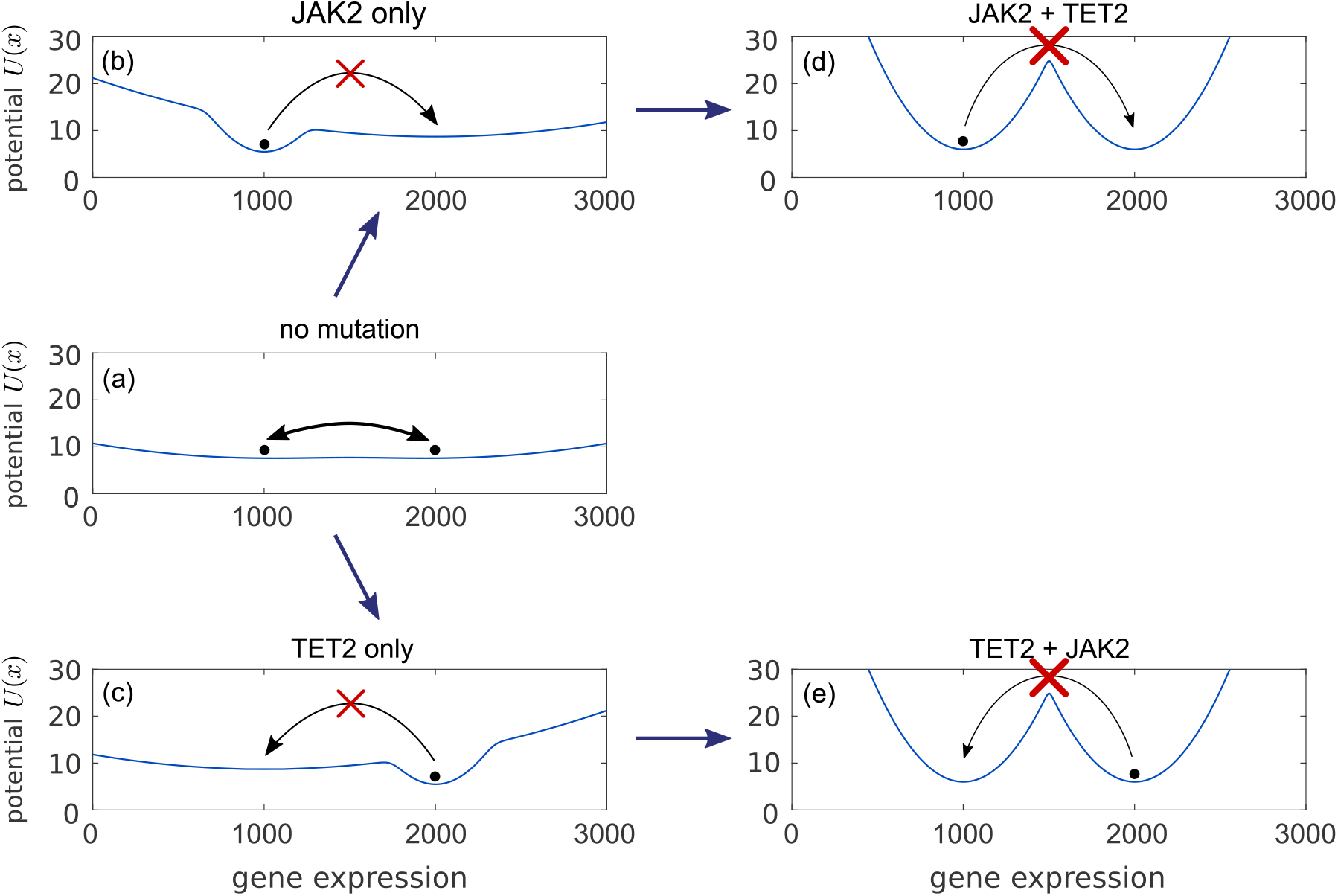
The effective potential function *U* (*x*) of gene expression levels *x* for different scenarios within the Markov chain model. (a) The potential function in the absence of mutations. The system can switch between the shallow wells near *x* = 1000 and *x* = 2000. (b) *U* (*x*) when only the JAK2 mutation is present. The system is confined to the deep well near *x* = 1000. (c) *U* (*x*) when only the TET2 mutation is present. The system is confined to the deep well near the higher expression level *x* = 2000. (d) The potential function of X gene expression if the TET2 mutation appeared after the JAK2 mutation. Since the system was previously confined in the well near *x* = 1000, it will retain this lower level of expression for a long time. *U* (*x*) for a cell which acquired the JAK2 mutation from a mother cell which already had the TET2 mutation. Since the system was previously confined in the well near *x* = 2000, its gene expression will stay high for a long time. Theoretically, in the (unrealistically) long time limit, the probability densities in both cases (d) and (e) will equipartition itself symmetrically across the two equally deep wells.

In this model, different orders of mutations (JT and TJ) lead to the same final stationary distribution (Fig. 6(f)). However, different histories lead to concentration of probabilities to different wells (Figs. 6(d) and (e)) on finite timescales. If this mesoscopic time scale is comparable to the life span of human being, then the final stationary distribution is *de facto* inaccessible. This simple probabilistic model can also explain the difference in gene expression levels between patients with different orders of mutations.

## References

[1] Altrock, P. M., Liu, L. L., and Michor, F. The mathematics of cancer: integrating quantitative models. Nature Reviews Cancer 15, 12 (2015), 730–745.

[2] Angelini, E., Wang, Y., Zhou, J. X., Qian, H., and Huang, S. A model for the intrinsic limit of cancer therapy: Duality of treatment-induced cell death and treatment-induced stemness. PLoS Computational Biology 18, 7 (2022), e1010319.

[3] Ascolani, G., and Lio, P. Modeling breast cancer progression to bone: how driver mutation order and metabolism matter. BMC Medical Genomics 12 (2019), 1–19.

[4] Baik, R., Wyman, S. K., Kabir, S., and Corn, J. E. Genome editing to model and reverse a prevalent mutation associated with myeloproliferative neoplasms. PLoS One 16, 3 (2021), e0247858.

[5] Benferhat, S., and Smaoui, S. Possibilistic causal networks for handling interventions: A new propagation algorithm. In Proceedings of the National Conference on Artificial Intelligence (2007), vol. 22, Menlo Park, CA; Cambridge, MA; London; AAAI Press; MIT Press; 1999, p. 373.

[6] Bocci, F., Zhou, P., and Nie, Q. spliceJAC: transition genes and state-specific gene regulation from single-cell transcriptome data. Molecular Systems Biology 18, 11 (2022), e11176.

[7] Caravagna, G., Giarratano, Y., Ramazzotti, D., Tomlinson, I., Graham, T. A., Sanguinetti, G., and Sottoriva, A. Detecting repeated cancer evolution from multi-region tumor sequencing data. Nature Methods 15, 9 (2018), 707–714.

[8] Chen, X., Wang, Y., Feng, T., Yi, M., Zhang, X., and Zhou, D. The overshoot and phenotypic equilibrium in characterizing cancer dynamics of reversible phenotypic plasticity. Journal of Theoretical Biology 390 (2016), 40–49.

[9] Cheng, X., D’Orsogna, M. R., and Chou, T. Mathematical modeling of depressive disorders: Circadian driving, bistability and dynamical transitions. Computational and Structural Biotechnology Journal 19 (2021), 664–690.

[10] Chiba, S. Dysregulation of tet2 in hematologic malignancies. International Journal of Hematology 105 (2017), 17–22.

[11] Chou, T., and Wang, Y. Fixation times in differentiation and evolution in the presence of bottlenecks, deserts, and oases. Journal of Theoretical Biology 372 (2015), 65–73.

[12] Clarke, M. A., Woodhouse, S., Piterman, N., Hall, B. A., and Fisher, J. Using state space exploration to determine how gene regulatory networks constrain mutation order in cancer evolution. Automated reasoning for systems biology and medicine (2019), 133–153.

[13] Dawson, M. A., Bannister, A. J., Göttgens, B., Foster, S. D., Bartke, T., Green, A. R., and Kouzarides, T. JAK2 phosphorylates histone H3Y41 and excludes HP1α from chromatin. Nature 461, 7265 (2009), 819–822.

[14] De Bie, J., Demeyer, S., Alberti-Servera, L., Geerdens, E., Segers, H., Broux, M., De Keersmaecker, K., Michaux, L., Vandenberghe, P., Voet, T., et al. Single-cell sequencing reveals the origin and the order of mutation acquisition in T-cell acute lymphoblastic leukemia. Leukemia 32, 6 (2018), 1358–1369.

[15] Delhommeau, F., Dupont, S., Valle, V. D., James, C., Trannoy, S., Massé, A., Kosmider, O., Le Couedic, J.-P., Robert, F., Alberdi, A., et al. Mutation in TET2 in myeloid cancers. New England Journal of Medicine 360, 22 (2009), 2289–2301.

[16] Fudenberg, D., Imhof, L., Nowak, M. A., and Taylor, C. Stochastic evolution as a generalized moran process. Unpublished manuscript 15 (2004).

[17] Gao, Y., Gaither, J., Chifman, J., and Kubatko, L. A phylogenetic approach to inferring the order in which mutations arise during cancer progression. PLoS Computational Biology 18, 12 (2022), e1010560.

[18] Garcia, V., Bonhoeffer, S., and Fu, F. Cancer-induced immunosuppression can enable effectiveness of immunotherapy through bistability generation: A mathematical and computational examination. Journal of Theoretical Biology 492 (2020), 110185.

[19] Hamanishi, J., Mandai, M., Iwasaki, M., Okazaki, T., Tanaka, Y., Yamaguchi, K., Higuchi, T., Yagi, H., Takakura, K., Minato, N., et al. Programmed cell death 1 ligand 1 and tumor-infiltrating CD8+ T lymphocytes are prognostic factors of human ovarian cancer. Proceedings of the National Academy of Sciences 104, 9 (2007), 3360–3365.

[20] Hanahan, D., and Weinberg, R. A. Hallmarks of cancer: the next generation. Cell 144, 5 (2011), 646–674.

[21] Herbet, M., Salomon, A., Feige, J.-J., and Thomas, M. Acquisition order of Ras and p53 gene alterations defines distinct adrenocortical tumor phenotypes. PLoS Genetics 8, 5 (2012), e1002700.

[22] Ito, S., DAlessio, A. C., Taranova, O. V., Hong, K., Sowers, L. C., and Zhang, Y. Role of Tet proteins in 5mC to 5hmC conversion, ES-cell self-renewal and inner cell mass specification. Nature 466, 7310 (2010), 1129–1133.

[23] Jiang, D.-Q., Wang, Y., and Zhou, D. Phenotypic equilibrium as probabilistic convergence in multi-phenotype cell population dynamics. PLoS One 12, 2 (2017), e0170916.

[24] Kent, D. G., and Green, A. R. Order matters: the order of somatic mutations influences cancer evolution. Cold Spring Harb. Perspect. Med. 7, 4 (2017), a027060.

[25] Khakabimamaghani, S., Ding, D., Snow, O., and Ester, M. Uncovering the subtype-specific temporal order of cancer pathway dysregulation. PLoS Computational Biology 15, 11 (2019), e1007451.

[26] Kim, L. U., D’Orsogna, M. R., and Chou, T. Perturbing the hypothalamicpituitaryadrenal axis: A mathematical model for interpreting ptsd assessment tests. Computational Psychiatry (Feb 2018).

[27] Klampfl, T., Gisslinger, H., Harutyunyan, A. S., Nivarthi, H., Rumi, E., Milosevic, J. D., Them, N. C., Berg, T., Gisslinger, B., Pietra, D., et al. Somatic mutations of calreticulin in myeloproliferative neoplasms. New England Journal of Medicine 369, 25 (2013), 2379–2390.

[28] Koushyar, S., Economides, G., Zaat, S., Jiang, W., Bevan, C. L., and Dart, D. The prohibitin-repressive interaction with E2F1 is rapidly inhibited by androgen signalling in prostate cancer cells. Oncogenesis 6, 5 (2017), e333–e333.

[29] Levine, A. J., Jenkins, N. A., and Copeland, N. G. The roles of initiating truncal mutations in human cancers: the order of mutations and tumor cell type matters. Cancer Cell 35, 1 (2019), 10–15.

[30] Levine, R. L., and Gilliland, D. G. JAK-2 mutations and their relevance to myeloproliferative disease. Current Opinion in Hematology 14, 1 (2007), 43–47.

[31] Li, X., and Levine, H. Bistability of the cytokine-immune cell network in a cancer microenvironment. Convergent Science Physical Oncology 3, 2 (2017), 024002.

[32] Lynch, M. Evolution of the mutation rate. TRENDS in Genetics 26, 8 (2010), 345–352.

[33] Mazaya, M., Trinh, H.-C., and Kwon, Y.-K. Effects of ordered mutations on dynamics in signaling networks. BMC Medical Genomics 13 (2020), 1–12.

[34] Mellman, I., Coukos, G., and Dranoff, G. Cancer immunotherapy comes of age. Nature 480, 7378 (2011), 480–489.

[35] Nakatake, M., Monte-Mor, B., Debili, N., Casadevall, N., Ribrag, V., Solary, E., Vainchenker, W., and Plo, I. JAK2V617F negatively regulates p53 stabilization by enhancing MDM2 via La expression in myeloproliferative neoplasms. Oncogene 31, 10 (2012), 1323–1333.

[36] Nangalia, J., Massie, C. E., Baxter, E. J., Nice, F. L., Gundem, G., Wedge, D. C., Avezov, E., Li, J., Kollmann, K., Kent, D. G., et al. Somatic CALR mutations in myeloproliferative neoplasms with nonmutated JAK2. New England Journal of Medicine 369, 25 (2013), 2391–2405.

[37] Nangalia, J., Nice, F. L., Wedge, D. C., Godfrey, A. L., Grinfeld, J., Thakker, C., Massie, C. E., Baxter, J., Sewell, D., Silber, Y., et al. Dnmt3a mutations occur early or late in patients with myeloproliferative neoplasms and mutation order influences phenotype. Haematologica 100, 11 (2015), e438.

[38] Niu, Y., Wang, Y., and Zhou, D. The phenotypic equilibrium of cancer cells: From average-level stability to path-wise convergence. Journal of Theoretical Biology 386 (2015), 7–17.

[39] Ohtani, K., Iwanaga, R., Nakamura, M., Ikeda, M.-a., Yabuta, N., Tsuruga, H., and Nojima, H. Cell growth-regulated expression of mammalian MCM5 and MCM6 genes mediated by the transcription factor E2F. Oncogene 18, 14 (1999), 2299–2309.

[40] Ortmann, C. A., Kent, D. G., Nangalia, J., Silber, Y., Wedge, D. C., Grinfeld, J., Baxter, E. J., Massie, C. E., Papaemmanuil, E., Menon, S., et al. Effect of mutation order on myeloproliferative neoplasms. N. Engl. J. Med. 372, 7 (2015), 601–612.

[41] Pastore, F., Bhagwat, N., Pastore, A., Radzisheuskaya, A., Karzai, A., Krishnan, A., Li, B., Bowman, R. L., Xiao, W., Viny, A. D., et al. PRMT5 Inhibition Modulates E2F1 Methylation and Gene-Regulatory Networks Leading to Therapeutic Efficacy in JAK2V617F-Mutant MPNPRMT5 Inhibition in MPN. Cancer Discovery 10, 11 (2020), 1742–1757.

[42] Pellegrina, L., and Vandin, F. Discovering significant evolutionary trajectories in cancer phylogenies. Bioinformatics 38, Supplement 2 (2022), ii49–ii55.

[43] Quan, J., and Wang, X.-J. Evolutionary games in a generalized moran process with arbitrary selection strength and mutation. Chinese Physics B 20, 3 (2011), 030203.

[44] Ramazzotti, D., Graudenzi, A., De Sano, L., Antoniotti, M., and Caravagna, G. Learning mutational graphs of individual tumour evolution from single-cell and multi-region sequencing data. BMC Bioinformatics 20, 1 (2019), 1–13.

[45] Reyes, M. E., Ma, J., Grove, M. L., Ater, J. L., Morrison, A. C., and Hildebrandt, M. A. RNA sequence analysis of inducible pluripotent stem cell-derived cardiomyocytes reveals altered expression of DNA damage and cell cycle genes in response to doxorubicin. Toxicology and Applied Pharmacology 356 (2018), 44–53.

[46] Roquet, N., Soleimany, A. P., Ferris, A. C., Aaronson, S., and Lu, T. K. Synthetic recombinase-based state machines in living cells. Science 353, 6297 (2016), aad8559.

[47] Shih, A. H., Abdel-Wahab, O., Patel, J. P., and Levine, R. L. The role of mutations in epigenetic regulators in myeloid malignancies. Nature Reviews Cancer 12, 9 (2012), 599–612.

[48] Smart, M., Goyal, S., and Zilman, A. Roles of phenotypic heterogeneity and microenvironment feedback in early tumor development. Phys. Rev. E 103 (2021), 032407.

[49] Swanton, C. Cancer evolution constrained by mutation order. New England Journal of Medicine 372, 7 (2015), 661–663.

[50] Talarmain, L. Modelling timing in blood cancers. Ph.D. thesis, University of Cambridge, 2021.

[51] Talarmain, L., Clarke, M. A., Shorthouse, D., Cabrera-Cosme, L., Kent, D. G., Fisher, J., and Hall, B. A. HOXA9 has the hallmarks of a biological switch with implications in blood cancers. Nature Communications 13, 1 (2022), 5829.

[52] Teimouri, H., and Kolomeisky, A. B. Temporal order of mutations influences cancer initiation dynamics. Physical Biology 18, 5 (2021), 056002.

[53] Turajlic, S., Xu, H., Litchfield, K., Rowan, A., Horswell, S., Chambers, T., O’Brien, T., Lopez, J. I., Watkins, T. B., Nicol, D., et al. Deterministic evolutionary trajectories influence primary tumor growth: TRACERx renal. Cell 173, 3 (2018), 595–610.

[54] Vithanage, G., Wei, H.-C., and Jang, S. R. Bistability in a model of tumor-immune system interactions with an oncolytic viral therapy. Apoptosis 1 (2021), 7.

[55] Wang, Y., and He, S. Inference on autoregulation in gene expression with variance-to-mean ratio. Journal of Mathematical Biology 86, 5 (2023), 87.

[56] Wang, Y., and Qian, H. Mathematical representation of Clausius’ and Kelvin’s statements of the second law and irreversibility. Journal of Statistical Physics 179, 3 (2020), 808–837.

[57] Wang, Y., and Wang, Z. Inference on the structure of gene regulatory networks. Journal of Theoretical Biology 539 (2022), 111055.

[58] Yu, Z., Sun, Y., She, X., Wang, Z., Chen, S., Deng, Z., Zhang, Y., Liu, Q., Liu, Q., Zhao, C., et al. SIX3, a tumor suppressor, inhibits astrocytoma tumorigenesis by transcriptional repression of AURKA/B. Journal of Hematology & Oncology 10 (2017), 1–16.

[59] Zhou, D., Wang, Y., and Wu, B. A multi-phenotypic cancer model with cell plasticity. Journal of Theoretical Biology 357 (2014), 35–45.

